# Telemedicine’s Impact on Diabetes Care during the COVID-19 Pandemic: A Cohort Study in a Large Integrated Healthcare System

**DOI:** 10.1101/2024.02.25.24303335

**Authors:** Reysha Patel, Jie Huang, Loretta Hsueh, Anjali Gopalan, Andrea Millman, Isabelle Franklin, Mary Reed

## Abstract

**Introduction:** To examine if patients exposed to primary care telemedicine (telephone or video) early in the COVID-19 pandemic had higher rates of downstream HbA_1c_ measurement and improved HbA_1c_ levels in the second year of the pandemic.

**Research Design and Methods:** In a cohort of 242, 848 Kaiser Permanente Northern California patients with diabetes, we examined associations between early-pandemic patient-initiated telemedicine visit and downstream HbA_1c_ monitoring and results during the second year of the pandemic.

**Results:** Adjusted HbA_1c_ measurement rates were significantly higher among patients with telemedicine exposure in the early-pandemic prior year than those with no visits in the prior year (91.0% testing for patients with video visits, 90.5% for telephone visits, visits, 86.7% for no visits, *p* < 0.05). Among those with HbA_1c_ measured, the rates of having an HbA_1c_ < 8% in the second year of the COVID-19 pandemic were also statistically significantly higher among patients with telemedicine exposure in the early-pandemic prior year than those with no visits in the prior year (68.5% with HbA_1c_< 8% for video visits, 67.3% for telephone visits, 66.6% for no visits, *p* < 0.05).

**Conclusions:** Access to telephone and video telemedicine throughout the early COVID-19 pandemic was associated with patients’ continued engagement in recommended diabetes care. Although our study analyzed telemedicine use during a pandemic, telemedicine visits may continue to support ongoing health care access and positive clinical outcomes.

**KEY MESSAGES:** The pandemic significantly increased telemedicine adoption, providing an opportunity to maintain health care access for patients with diabetes. Our study investigates the impact of telemedicine, including both telephone and video visits, on diabetes care during the early COVID-19 period. The results demonstrate that patients utilizing telemedicine exhibit higher rates of HbA_1c_ measurement and at-goal HbA_1c_. These findings suggest that telemedicine can be a valuable tool in supporting clinical outcomes in the management of chronic health conditions.

## INTRODUCTION

Prior to the COVID-19 pandemic and its associated need for shelter-in-place orders, telemedicine utilization was low, with nearly 80% of U.S. physicians reporting no prior telemedicine experience.^1^ During shelter-in-place, telemedicine use expanded rapidly and emerged as a tool in supporting continued healthcare access while maintaining physical distancing measures, including for primary care delivery and chronic condition management.^2,3,4^ This growth was aided by public policy changes that temporarily loosened restrictions on telehealth services and increased reimbursement for virtual visits.^5, 6^ Though in-person visits are now returning, the laws that allowed for broader access to telehealth services have largely stayed in place and telemedicine is continuing to be practiced at a high rate, indicating that the change brought on by COVID-19 is likely to have lasting effects on healthcare delivery.^7, 8^ Research is needed to assess the impact of telemedicine expansion on the care and health outcomes of patients with chronic diseases.

Over 35 million Americans have diabetes^9^ and its management can be complex, requiring ongoing monitoring and adjustment by clinicians. Even prior to the COVID-19 pandemic, studies were being conducted on the efficacy of telemedicine in diabetes management. Most showed telemedicine to be at least as effective as standard care when evaluating outcomes such as HbA_1c_.^10, 11, 12, 13, 14, 15, 16^ Research since the onset of the pandemic continues to demonstrate the efficacy of telemedicine in diabetes management; however, these studies are primary RCTs which may not accurately represent real-world clinical practice and do not consider the pandemic-driven increase in telemedicine adoption.^17, 18,19^ Early studies examining telemedicine uptake during the COVID-19 pandemic offer promising short-term results.^20, 21^ Nonetheless, evaluating telemedicine’s impact on health outcomes, particularly in chronic conditions such as diabetes, beyond the initial pandemic phase remains essential. This knowledge will play an important role in shaping continued telemedicine access and reimbursement, which will particularly impact diabetes patients in need of consistent primary care.

This study seeks to understand the association between telemedicine visits during the early pandemic and diabetes management in the subsequent year, including HbA_1c_ testing and rates of HbA_1c_ levels < 8%. To examine patients based on their baseline pre-pandemic diabetes severity, we stratified patients based on baseline pre-pandemic HbA_1c_ level (<8% or 8+%) before the pandemic. We hypothesize that patient health care access through patient-initiated primary care telemedicine visits (telephone or video) early in the COVID-19 pandemic would support higher rates of HbA_1c_ measurement and improved HbA_1c_ levels compared to those without any visit with their primary care provider (PCP) during the early-pandemic.

## RESEARCH DESIGN AND METHODS

### Setting

In this cohort study conducted at Kaiser Permanente Northern California (KPNC), a large integrated health care delivery system, we examined associations between patient-initiated telemedicine PCP visits in the first year of the COVID-19 pandemic and downstream HbA_1c_ management during the second year of the COVID-19 pandemic. As in an office primary care visit, clinicians conducting telemedicine visits had full access to the patient’s electronic health record (EHR) history and documented telemedicine visits directly within the EHR. Patients could receive a call for a telephone visit on any phone number and could access video visits from home or elsewhere through any internet-connected and video-enabled computer or mobile device.

### Study Design and Population

We studied all patients included in the KPNC clinical diabetes registry who maintained continuous health plan membership throughout the three-year study period. The study period was divided into three distinct time periods to facilitate analysis. The baseline period (04/2019-03/2020) encompassed the year preceding the COVID-19 pandemic and was used to stratify based on patients’ pre-pandemic HbA_1c_ levels. The telemedicine exposure period (04/2020-03/2021) represented the first year of the pandemic when various state and federal shelter-in-place measures were implemented to mitigate the spread of COVID-19. During this period, all patient-initiated primary care visits at KPNC were restricted to telemedicine first (video or telephone visits) and any subsequent in-person visits were at the discretion of the providers. The diabetes outcome period (04/2021-03/2022) was the second year of the pandemic, characterized by fewer pandemic restrictions and policies, and full access to patient-initiated in-person primary care visits. We assessed diabetes outcomes in this second year of the pandemic, among patients exposed to telemedicine in the prior year (compared with those who were not exposed to telemedicine).

### Exposure and Outcome Measures

To assess the impact of telemedicine exposure, we categorized patients based on their primary care visit utilization during the first year of the pandemic: any video visit (with or without any telephone visit), telephone visits only (without any video visit), or no patient-initiated primary care visits. Our objective was to determine whether having a patient-initiated telemedicine primary care visit during the exposure period was associated with two diabetes care outcomes: (1) any HbA_1c_ measurement during the diabetes outcome period, and (2) among those who had any measurement, an HbA_1c_ < 8% (first measurement during the outcome period).

### Covariates

In comparing outcomes related to telemedicine utilization, we accounted for an extensive range of covariates. The model adjusted for patient sociodemographic factors (patient age, sex, race/ethnicity, neighborhood socioeconomic status, neighborhood internet access, English language preference), technology access factors (history of mobile access to a patient portal, history of a video visit during the baseline period, family member proxy access to the patient’s online portal), patient clinical characteristics (presence of another chronic condition, history of a flu shot, baseline HbA_1c_ measurement, medical center, and the month when HbA_1c_ was measured). These covariates were selected to address factors such as access to technology and the propensity to seek healthcare services. We used the EHR and other administrative data sources to identify patient sociodemographic characteristics (age, sex, race and ethnicity, and language preference), patient’s residential address to define patient neighborhood socioeconomic status (2010 US census measures at the census block group level), and neighborhood residential high-speed internet access level (Federal Communications Commission census tract level data). **Statistical Analysis**

We employed multivariable logistic regression to examine the association between exposure to primary care telemedicine during the telemedicine exposure period with each of the study diabetes outcomes during the diabetes outcome period (any HbA_1c_ measurement, or HbA_1c_ < 8%). The analysis was repeated by stratifying patients based on their HbA_1c_ level (< 8% or 8% +) during the pre-pandemic baseline period as a measure of clinical need prior to the COVID-19 pandemic. Those without an HbA_1c_ measurement during the outcome period were excluded from the analysis of HbA1c level. In instances where multiple HbA_1c_ measurements were taken during the diabetes outcome period, the initial measurement was used to assess HbA_1c_ management.

For ease of interpretation, we calculated adjusted percentages by marginal standardization using post-estimation command ‘margins’ via Stata 17.0. All analyses were conducted using 2-sided tests for significance at *p*□<□0.05.

## RESULTS

Among 242,848 individuals with diabetes in the study sample, 42.9% had a primary care video visit (with or without a phone visit), 32.3% had only primary care telephone visits, and 25.0% had no primary care visits during the first year of the COVID-19 pandemic (telemedicine exposure period). Overall, 60.0% of the patients were non-White, 51.7% were male, 41.0% were between the ages of 45 and 64, 87.4% preferred English, 23.7% were from low socioeconomic status neighborhoods, 30.6% had low neighborhood internet, and 69.0% had a baseline HbA_1c_ < 8% (Table 1).

**Table 1.**
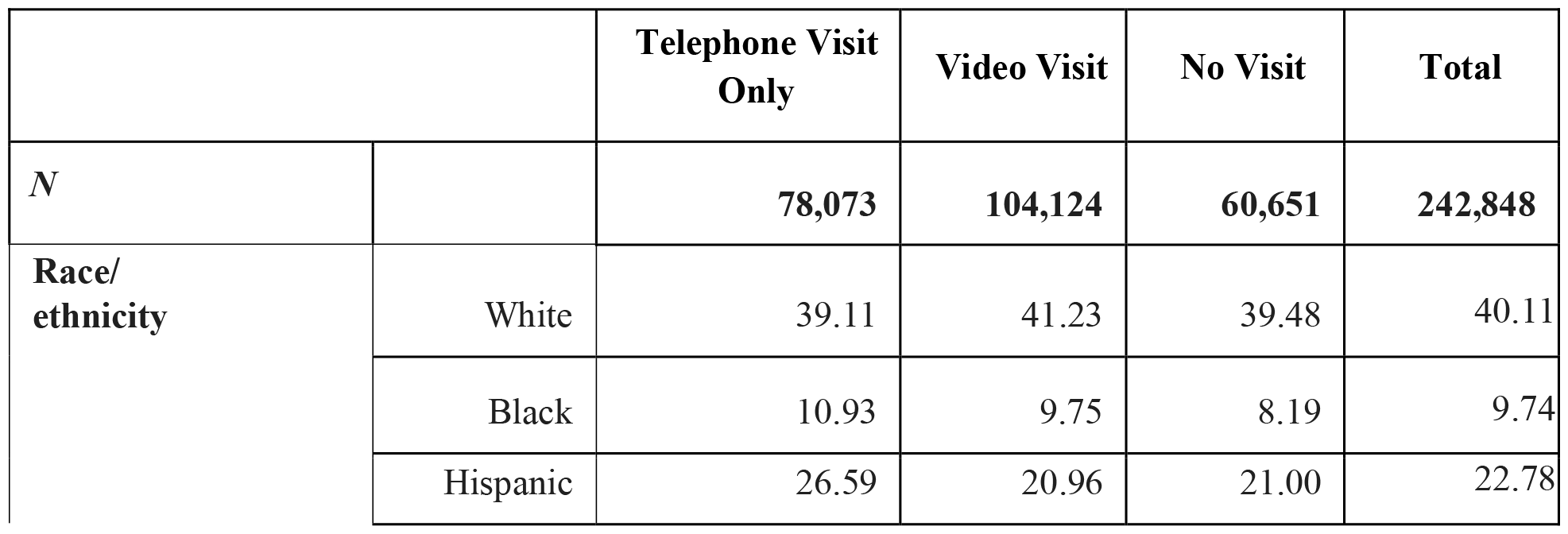
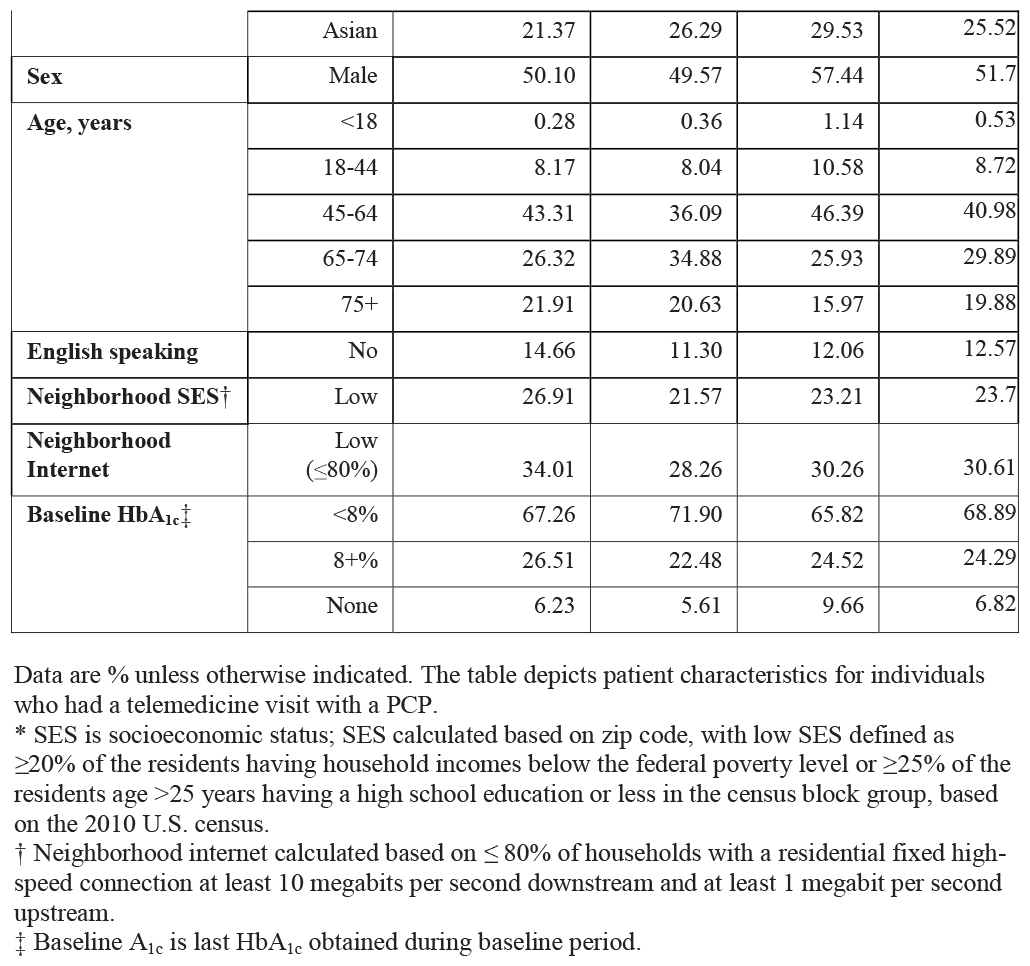
Patient characteristics for individuals who had a telemedicine visit with a PCP.

After adjustment, the rates of having an HbA_1c_ measurement during the second year of the COVID-19 pandemic (diabetes outcome period) were statistically significantly higher among patients with telemedicine exposure than those with no visits (91.0% for video visits, 90.5% for telephone visits, visits, 86.7% for no visits, *p* < 0.05; Figure 1). Among those with an HbA_1c_ measured, the proportion with an HbA_1c_ < 8% in the second year of the COVID-19 pandemic was also statistically significantly higher among people with telemedicine exposure in the prior year than those with no visits in the prior year (68.5% for video visits, 67.3% for telephone visits, 66.6% for no visits, *p* < 0.05; Figure 2).

**Figure 1.**
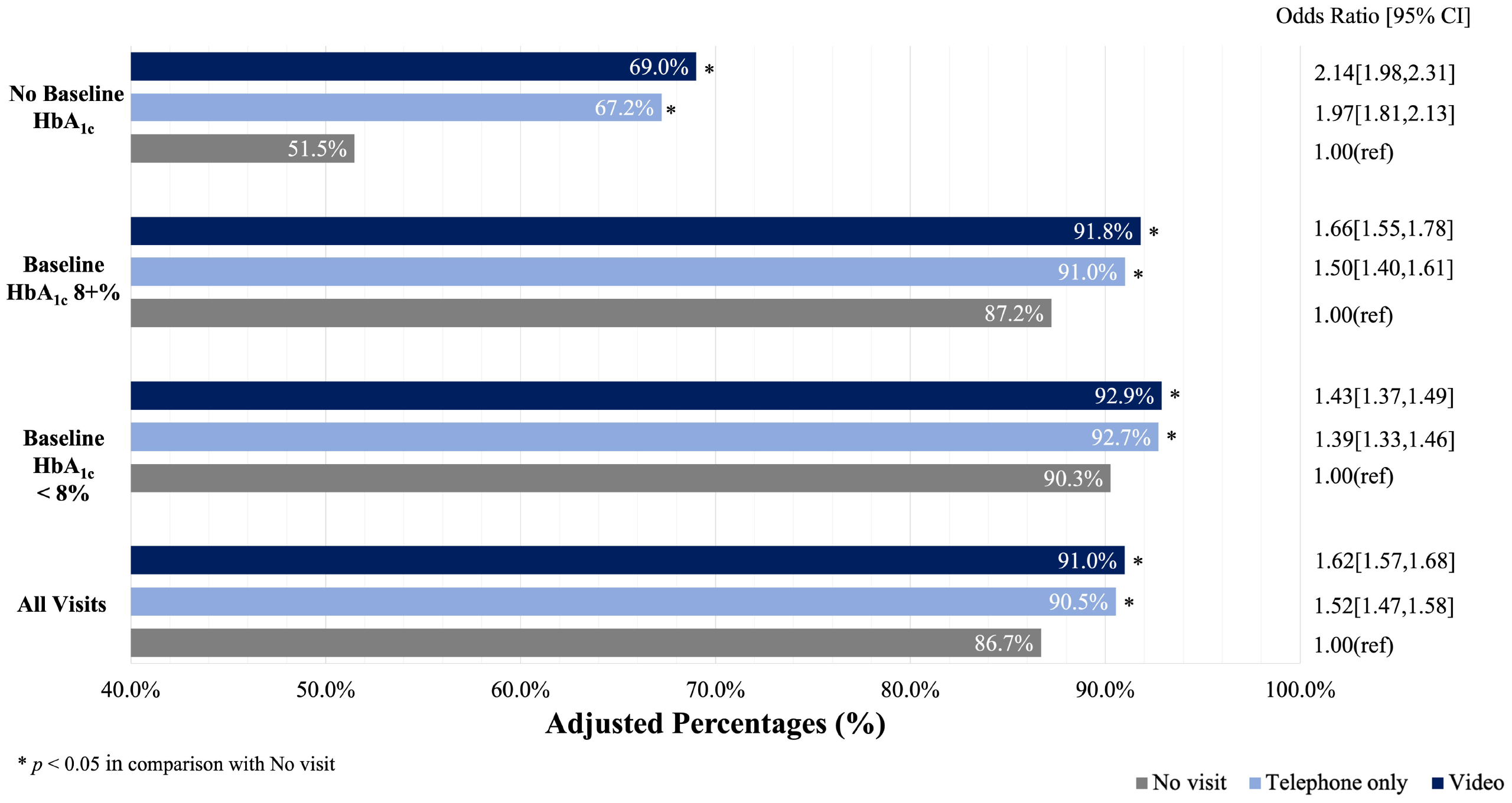
Adjusted association of pandemic telemedicine PCP visit with any HbA_1c_ measurement. Rates of having an HbA_1c_ measurement during the second year of the COVID-19 pandemic were statistically significantly higher among patients with telemedicine exposure than those with no visits (91.0% for video visits, 90.5% for telephone visits, visits, 86.7% for no visits, *p* < 0.05).

**Figure 2.**
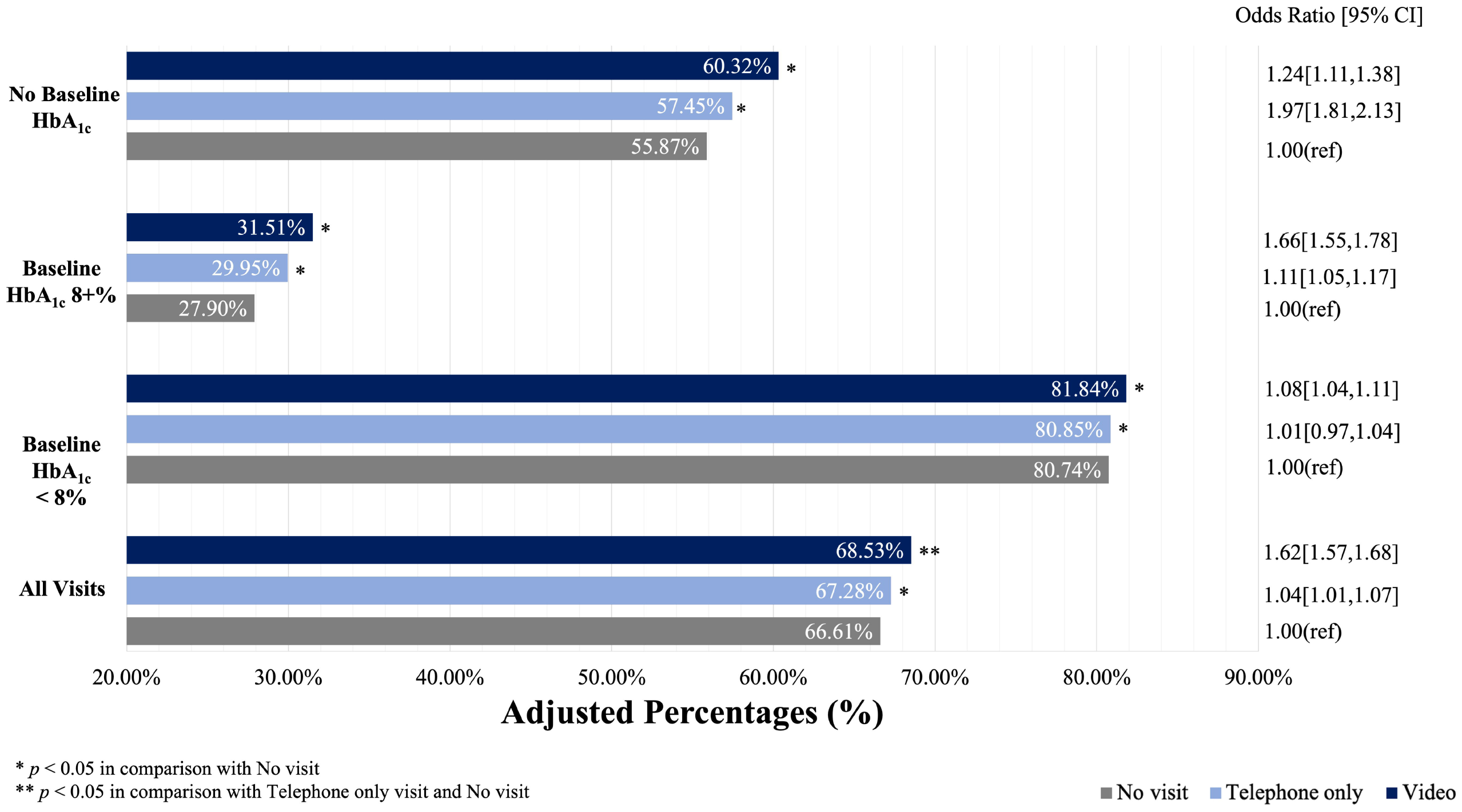
Adjusted association of pandemic telemedicine PCP visit with a HbA_1c_ < 8%. Among those with an HbA_1c_ measured, the proportion with an HbA1c < 8% in the second year of the COVID-19 pandemic was statistically significantly higher among people with telemedicine exposure in the prior year than those with no visits in the prior year (68.5% for video visits, 67.3% for telephone visits, 66.6% for no visits, *p* < 0.05).

While the direction and statistical significance of these results were consistent among all groups when stratifying patients based on baseline HbA_1c_, telemedicine-associated outcome rates were stronger among patients with HbA_1c_ of 8%+ at baseline than those already with HbA_1c_ < 8% prior to the pandemic. Further, among patients with no HbA_1c_ measured in the baseline pre-pandemic year, exposure to telemedicine in the first pandemic year was associated with the largest marginal benefit (17.5% higher HbA_1c_ measurement among patients exposed to video visits than patients without any primary care visits, and 4.5% higher rates of HbA_1c_ < 8% among patients exposed to video visits than patients without any primary care visits). Rates of HbA_1c_ control were also statistically significantly higher among patients with video visits than those with telephone-only visits in the prior year.

## CONCLUSIONS

In a study of patient-initiated primary care telemedicine use during the early COVID-19 pandemic in a large integrated delivery system, access to telephone and video telemedicine during a period of limited access to in-person visits supported diabetes care and outcomes.

Although our study analyzed telemedicine use during a pandemic, our results show that telemedicine visits are associated with positive clinical outcomes for patients with diabetes. Regular visits and adherence to multiple preventative measures are key to preventing diabetes-related long-term health complications.^22^ Telemedicine offers the opportunity for more frequent interactions for education and counseling, which are associated with improved health outcomes in people with diabetes.^23^ Additionally, it facilitates patient and PCP communication while reducing entry barriers, addressing challenges like transportation and time constraints.^24^

The study results also suggested that video visits might offer more comprehensive care and higher rates of diabetes control compared to telephone visits. Previous studies have shown that this is potentially due to the ability to engage in nonverbal communication and build a stronger patient-provider relationship over video.^25, 26^ Moving forward, efforts to ensure access for all patients should be prioritized, especially since rates of telemedicine use remain high since the pandemic.^7, 8^ Future policy determining the fate of long-term reimbursement for telemedicine visits should account for their potential to improve health outcomes and broaden the availability of healthcare services.

This study includes limitations. While our analyses accounted for a wide range of patient and clinical covariates, we could not rule out the omission of unmeasured confounding variables due to the observational nature of the study. As a result, causal conclusions cannot be drawn from the findings. For example, individuals who are more actively engaged in their health may be more inclined to have telemedicine visits, and these patients are also more likely to have an HbA_1c_ measurement and well managed diabetes, even in the absence of a visit. Furthermore, the data analyzed in this study was collected during a specific study period amidst the COVID-19 pandemic. The unique circumstances and restrictions during this period may limit the generalizability of the data to post-pandemic telemedicine use. Further research on telemedicine use beyond the COVID-19 pandemic is needed, especially as the dynamics between telemedicine and in-person services, and telephone and video visits, continue to evolve. Additionally, the study was conducted in a single healthcare setting with longstanding telemedicine use, which may not generalize to settings with less extensive experience with telemedicine. The study design aimed to assess the association between primary care visits during the study period, however, we do not measure or account for other health care factors during the study period that might impact diabetes outcomes. Finally, our study design is not able to examine patient outcomes in any patient who did not have any HbA_1c_ measurement during the outcomes period.

Primary care telemedicine visits can provide patients with access to high-quality remote healthcare, making it a valuable tool, especially in cases where in-person communication is challenging or not feasible. Telemedicine might be especially valuable for patients who live in rural communities or face transportation barriers. In a study of the association between telemedicine visits and rates of HbA_1c_ measurement and management among patients with diabetes during the COVID-19 pandemic, telemedicine visits were associated with higher rates of HbA_1c_ measurement and higher rates of HbA_1c_ < 8%, with particular benefits in patients whose HbA_1c_ was already < 8% before the pandemic started. Further, video visits were associated with additional benefits over telephone-only telemedicine visits. Still, further research is needed to fully understand the impact of telephone versus video visits on patient outcomes. Overall, telemedicine visits show promise as a valuable tool for connecting patients to healthcare providers and effectively managing chronic conditions, such as diabetes.

## Data Availability

All data produced in the present study are available upon reasonable request to the authors

## Funding and Assistance

This study was funded through NIDDK Medical Student Research Program, NIH/NIDDK T32DK116684, and R01 DK085070.

## Author Contributions and Guarantor Statement

M.R. and J.H. were involved in the conception, design, and conduct of the study and the analysis and interpretation of the results. R.P. wrote the first draft of the manuscript, and all authors edited, reviewed, and approved the final version of the manuscript. J.H. is the guarantor of this work and, as such, had full access to all the data in the study and takes responsibility for the integrity of the data and the accuracy of the data analysis.

## Ethics Statement

This study was reviewed and approved by the Kaiser Permanente Northern California IRB committee. The approval confirms that the study adheres to ethical guidelines and standards for research involving human subjects. The IRB protocol number associated with this approval is CN-04JHsu-02-H.

